# Young age, student status and reported non-binary gender associate strongly with decreased functioning during Covid-19 pandemic in a university community

**DOI:** 10.1101/2022.05.30.22275757

**Authors:** Raimo K. R. Salokangas, Tiina From, Jarmo Hietala

## Abstract

**Background:** Covid-19 pandemic has had detrimental effects on physical and mental well-being whereas there are fewer studies on Covid-19 effects on everyday functioning.

**Aims:** We aimed to investigate effects of Covid-19 on functioning and related factors in a university community.

**Method:** In all, 2004 students and university personnel responded to a Webropol survey in May 2021, when the measures for preventing Covid-19 infections had sustained about a year and a half. Functioning included Visual Analog Scale (0 to 10) assessments on ability to function and ability to work.

**Results:** Young age, reported non-binary gender, being student, low resilience, loneliness, received mental care and minor physical exercise, as well as depressive symptoms associated with inferior functioning and negative effects of Covid-19 on functioning. Good school performance at adolescence associated with better, while childhood adversities associated with poorer functioning.

**Conclusions:** In the university community, young age and non-binary gender associated with decreased functioning during Covid-19 pandemic. Functioning of students was lower than in that of the university personnel. The need for therapeutic counselling and interventions is greatest among young students.

## Introduction

The Covid-19 global pandemic has had a negative impact on physical and mental health and well-being of many (Brooks et al., 2020); after the declaration of Covid-19, negative emotions (e.g., anxiety, depression, and indignation) and sensitivity to social risks increased, while positive emotions and life satisfaction decreased (Li et al., 2020).

Measures for preventing Covid-19 viral infections have resulted in various lockdowns of the society, like remote work and study, social distancing and home quarantine. Social isolation caused by these measures have increased subjective feeling of loneliness and likelihood of psychological distress, like depression and anxiety (Loades et al., 2020).

According to a meta-analysis, Covid-19 pandemic caused higher prevalence of anxiety and depression in study populations (Salari et al., 2020). However, detrimental effects of Covid-19 do not distribute evenly in the society. In population surveys young age, female and non-binary gender, working outside home, living with children and low resilience and assets have associated with psychological distress experiences, like depression, anxiety and post-traumatic stress symptoms (Gómez-Salgado et al., 2020; Prout et al., 2020; Guldager et al., 2021; Hou et al., 2021a, 2021b; Schmits et al., 2021).

Ability to response to acute adversities, like Covid-19, depends also on individual’s resilience (Prati & Mancini, 2021). During the first weeks of the nation-wide lockdown efforts, average resilience was lower than expected. However, it was greater among those who tended to get outside more often, exercise more, perceive more social support from family, friends, and significant others (Killgore et al., 2020). Among younger students, emotional resilience was positively correlated with learning management skills that were predicted by positive emotional ability (Zhang et al., 2020b).

Especially students have suffered from negative effects of Covid-19 pandemic. In a study of university students, 71% indicated increased stress and anxiety, 82% increased concerns on academic performance and 86% decreased social interactions due to the Covid-19 outbreak (Son et al., 2020). Students’ interaction and co-studying networks have become sparser; students have been more sedentary and reported increased loneliness, anxiety and depression (Elmer et al., 2020; Schmits et al., 2021). In a survey of medical and health science students, 39% reported academic stress due to Covid-19. Female sex, young age, bachelor level, knowing a Covid-19 patient and being worried about becoming infected were associated with academic stress (Guldager et al., 2021). A year after the beginning of the pandemic, many students in higher education were anxious and depressed. Contacts with family and friends as well as regular physical activity seemed to be protective factor against psychological distress (Schmits et al., 2021).

Studies related to psychosocial effects of Covid-19 pandemic have mainly focused on behavioural changes and mental distress whereas studies on the effect of Covid-19 specifically on everyday functioning are rare. In the present study, we aimed to investigate the effects of Covid-19 pandemic on individuals’ ability to function in a university community. We hypothesised that the social stress related to the Covid-19 pandemic may have had a specific damaging effect on functioning of younger students with a modifying effect of gender. Functioning of individuals, who have been sensitised to stress situations, like those who have faced adversities in childhood, and who currently have psychic symptoms may also be a specific group that has struggled during Covid-19 pandemic.

## Methods

The ethical committee of the University of Turku approved the study protocol. All study subjects gave their written consent to the study.

### Participants and procedure

In May 2021, a Webropol survey was sent to the personnel and students of the University of Turku. The survey dealt with study subject’s gender, background and living situation, adolescence school performance, friends and adversities, mental care, current contacts with family and friends, resilience, loneliness, psychic symptoms, current functioning (FUNCT) and subjective evaluation of the effect of Covid-19 on functioning (COVFUNCT).

The survey was mailed to 27784 subjects (21227 students and 6557 personnel members) and re-mailed a week after the first mailing. Study subjects responded anonymously. In all, 2004 returned the survey and 1998 subjects (7.4%) completed the whole inquiry. Number of missing data was about 1% or less.

### Questions

Gender question included four options 1) Female (Binary Female, BF), 2) Male (Binary Male, BM), 3) Other, 4) I do not wish to tell. In preliminary analyses on depressive and anxiety symptoms, and on functioning and effect of Covid-19 on functioning, the individuals, who selected two latest options separately, differed significantly from BF and BM individuals. In further analyses, two latter options were combined for indicating Non-Binary Gender (NBG). The combination of BF and BM was considered as Binary Gender (BG). For multivariate analyses, work status was re-classed into three classes: 1) employee (fulltime or part-time), 2) student (fulltime or part-time) and 3) other (at home, unemployed or sick).

Functioning was assessed with two questions: “How is your ability to function?” and “How is your ability to work?” using Visual Analog Scale (VAS) (0 = very bad … 10 = very good). Sum of these two questions divided by two indicated FUNCT. Correspondingly, two VAS questions: “How the corona epidemic affected to your ability to function/work?” (rated 0 = extremely negatively … 10 = extremely positively) were summed up and divided by 2 for indicating COVFUNCT.

School success and number of close friends at age of 12 to 18 were assessed with VAS (0 = very badly/no friends at all … 10 = very well/ a lot of close friends). Resilience was assessed with two VAS questions: 1) How quickly do you recover after an adversity? and 2) If you fail, how long does your failure bother you? (0 = very slowly/for a very long time … 10 = very quickly/for a very short period). Sum of these two questions divided by two was used as an indicator of resilience. Mental care was assessed with two questions: “Have you, during the last 6 months/ever, received care for mental health issues? (1 = Yes, 2 = No). Alcohol problems included three questions: 1)” I think that I use too much alcohol”, 2) “My friend or family member has said that I use too much alcohol”, 3) “I have sought professional help for my alcohol use and the problems it causes” (1 = Yes, 2 = No). Sum of these three questions indicated alcohol problems. Physical exercise was measured with the question: “How often do you exercise for at least half an hour at the time, so that you at least mildly get out of breath and sweat” (1 = daily, 2 = weekly, 3 = less often).

Number of contacts with family and close friends during a month were rated: 1 = none, 2 = one, 3 = two, 4 = three to four and 5 = five or more. Loneliness was a sum of three question: “How often do you feel that you 1) lack companionship, 2) are left out and 3) are isolated from others?” Each of question was rated: 1 = hardly ever, 2 = some of the time and 3 = often (Lubben et al., 2006). Childhood adversities were assessed with the Trauma and Distress Scale (Salokangas et al., 2016) which produces five core domains: Emotional, Physical and Sexual Abuse and Emotional and Physical Neglect. Depressive symptoms were assessed with the DEPS scale (Salokangas et al., 1995) and anxiety symptoms with BAI (Beck et al., 1988).

### Statistical analyses

Distributions of background characteristics were cross-tabulated with gender and tested with Khi-test (Table 1). Means of continuous variables were calculated by gender and tested in ANOVA (Table 2) and Pearson’s correlations were calculated for continuous variables (Supplementary table 1). In ANOVA, variance of FUNCT and COVFUNCT was explained by background characteristics (Table 1) and resilience, loneliness, family and friend contacts, alcohol problems and physical exercise, adolescence school success and friends and childhood adversities. The explanatory variables associating non-significantly with FUNCT/COVFUNCT were omitted one by one. In post hoc analyses, depressive and anxiety symptoms were entered into the final models.

**Table 1.** Functioning (FUNCT, 0-10) and effect of Covid-19 on functioning (COFUNCT, 0-10) by background characteristics.

|  |  | FUNCT |  |  | COVFUNCT |  |  |
| --- | --- | --- | --- | --- | --- | --- | --- |
|  | N | Mean | SD | p | Mean | SD | p |
| All | 1998 | 7.04 | 2.14 |  | 4.59 | 1.92 |  |
| Gender |  |  |  | <0.001 |  |  | <0.001 |
| Female | 1452 | 7.07 | 2.12 |  | 4.63 | 1.95 |  |
| Male | 475 | 7.21 | 2.13 |  | 4.67 | 1.83 |  |
| Other | 41 | 5.04 | 2.42 |  | 3.34 | 1.36 |  |
| Don't want to say | 30 | 6.13 | 1.84 |  | 3.47 | 1.50 |  |
| Age |  |  |  | <0.001 |  |  | <0.001 |
| -20 | 95 | 6.22 | 2.39 |  | 3.65 | 1.55 |  |
| 21-30 | 988 | 6.74 | 2.17 |  | 4.24 | 1.86 |  |
| 31-40 | 373 | 7.03 | 2.17 |  | 4.56 | 1.83 |  |
| 41-50 | 273 | 7.70 | 1.90 |  | 5.27 | 1.83 |  |
| 51-60 | 196 | 7.73 | 1.84 |  | 5.65 | 1.93 |  |
| 61-70 | 64 | 8.00 | 1.52 |  | 5.37 | 1.75 |  |
| 71+ | 9 | 7.44 | 2.08 |  | 5.67 | 1.95 |  |
| Marital status |  |  |  | <0.001 |  |  | <0.001 |
| Single | 875 | 6.65 | 2.17 |  | 4.22 | 1.81 |  |
| Married | 530 | 7.74 | 1.88 |  | 5.26 | 1.92 |  |
| Cohabitation | 516 | 6.97 | 2.15 |  | 4.48 | 1.90 |  |
| Divorced | 71 | 7.11 | 2.39 |  | 4.95 | 1.92 |  |
| Widowed | 6 | 7.75 | 1.84 |  | 4.92 | 0.66 |  |
| Living with |  |  |  | <0.001 |  |  | <0.001 |
| Alone | 777 | 6.67 | 2.20 |  | 4.21 | 1.80 |  |
| Parents | 47 | 6.50 | 2.45 |  | 4.57 | 1.95 |  |
| Spouse/children | 1048 | 7.37 | 2.06 |  | 4.91 | 1.97 |  |
| Someone else | 71 | 7.01 | 1.85 |  | 4.37 | 1.71 |  |
| Other living situation | 55 | 6.56 | 2.02 |  | 4.24 | 1.44 |  |
| Childhood family |  |  |  | <0.001 |  |  | 0.073 |
| Two parents | 1434 | 7.15 | 2.11 |  | 4.65 | 1.94 |  |
| One parent died | 70 | 7.34 | 1.81 |  | 4.80 | 2.12 |  |
| Parents divorced | 422 | 6.72 | 2.19 |  | 4.42 | 1.77 |  |
| Single parent | 31 | 6.52 | 2.87 |  | 4.24 | 1.96 |  |
| Other | 41 | 6.29 | 2.43 |  | 4.20 | 1.91 |  |
| Work status |  |  |  | <0.001 |  |  | <0.001 |
| Fulltime work | 715 | 7.75 | 1.73 |  | 5.17 | 1.85 |  |
| Part-time work | 69 | 7.06 | 1.98 |  | 4.45 | 2.15 |  |
| Family work | 12 | 7.83 | 2.31 |  | 4.67 | 1.50 |  |
| Fulltime study | 975 | 6.75 | 2.10 |  | 4.26 | 1.82 |  |
| Part-time study | 155 | 6.95 | 2.29 |  | 4.45 | 1.89 |  |
| Unemployed | 27 | 5.96 | 2.52 |  | 4.17 | 2.09 |  |
| Sick leave | 45 | 2.84 | 1.96 |  | 3.56 | 2.32 |  |
| Mental care; ever |  |  |  | <0.001 |  |  | <0.001 |
| Yes | 942 | 6.24 | 2.23 |  | 4.25 | 1.96 |  |
| No | 1056 | 7.75 | 1.78 |  | 4.90 | 1.82 |  |
| Mental care; past 6 months |  |  |  | <0.001 |  |  | <0.001 |
| Yes | 456 | 5.61 | 2.29 |  | 3.90 | 1.94 |  |
| No | 1542 | 7.47 | 1.90 |  | 4.80 | 1.86 |  |
| Alcohol problems |  |  |  | 0.007 |  |  | 0.358 |
| Yes | 201 | 6.66 | 2.31 |  | 4.48 | 1.99 |  |
| No | 1797 | 7.09 | 2.12 |  | 4.61 | 1.91 |  |
| Physical exercise |  |  |  | <0.001 |  |  | <0.001 |
| Daily | 131 | 7.83 | 1.92 |  | 5.44 | 2.24 |  |
| Weekly | 1446 | 7.21 | 2.05 |  | 4.62 | 1.86 |  |
| Less often | 421 | 6.22 | 2.31 |  | 4.24 | 1.92 |  |

**Table 2.** Pearson’s correlations

|  | 1. | 2. | 3. | 4. | 5. | 6. | 7. | 8. | 9. | 10. | 11. | 12. | 13. | 14. |
| --- | --- | --- | --- | --- | --- | --- | --- | --- | --- | --- | --- | --- | --- | --- |
| 1. Functioning | 1 |  |  |  |  |  |  |  |  |  |  |  |  |  |
| 2. Effect of Covid-19 on functioning | .554<sup>***</sup> | 1 |  |  |  |  |  |  |  |  |  |  |  |  |
| 3. Age | .206<sup>***</sup> | .276<sup>***</sup> | 1 |  |  |  |  |  |  |  |  |  |  |  |
| 4. Resilience | .418<sup>***</sup> | .281<sup>***</sup> | .231<sup>***</sup> | 1 |  |  |  |  |  |  |  |  |  |  |
| 5. Loneliness | -.419<sup>***</sup> | -.266<sup>***</sup> | -.235<sup>***</sup> | -.391<sup>***</sup> | 1 |  |  |  |  |  |  |  |  |  |
| 6. Family contacts | .177<sup>***</sup> | .103<sup>***</sup> | .088<sup>***</sup> | .135<sup>***</sup> | -.202<sup>***</sup> | 1 |  |  |  |  |  |  |  |  |
| 7. Friend contacts | .047<sup>*</sup> | -.061<sup>**</sup> | -.235<sup>***</sup> | .073<sup>**</sup> | -.271<sup>***</sup> | .105<sup>***</sup> | 1 |  |  |  |  |  |  |  |
| 8. School success (12-18) | .090<sup>**</sup> | -.004 | -.083<sup>***</sup> | .012 | -.063<sup>**</sup> | .087<sup>***</sup> | .109<sup>***</sup> | 1 |  |  |  |  |  |  |
| 9. Close friends (12-18) | .113<sup>***</sup> | .011 | .013 | .243<sup>***</sup> | -.269<sup>***</sup> | .146<sup>***</sup> | .313<sup>***</sup> | .176<sup>***</sup> | 1 |  |  |  |  |  |
| 10. Childhood adversities | -.295<sup>***</sup> | -.111<sup>***</sup> | -.011 | -.267<sup>***</sup> | .299<sup>***</sup> | -.266<sup>***</sup> | -.130<sup>***</sup> | -.182<sup>***</sup> | -.186<sup>***</sup> | 1 |  |  |  |  |
| 11. Depressive symptoms | -.702<sup>***</sup> | -.459<sup>***</sup> | -.258<sup>***</sup> | -.486<sup>***</sup> | .582<sup>***</sup> | -.216<sup>***</sup> | -.072<sup>**</sup> | -.057<sup>*</sup> | -.141<sup>***</sup> | .341<sup>***</sup> | 1 |  |  |  |
| 12. Anxiety symptoms | -.536<sup>***</sup> | -.322<sup>***</sup> | -.287<sup>***</sup> | -.436<sup>***</sup> | .439<sup>***</sup> | -.135<sup>***</sup> | -.010 | -.056<sup>*</sup> | -.087<sup>***</sup> | .354<sup>***</sup> | .673<sup>***</sup> | 1 |  |  |
| 13. Physical exercise | .208<sup>***</sup> | .134<sup>***</sup> | .051<sup>*</sup> | .125<sup>***</sup> | -.162<sup>***</sup> | .091<sup>***</sup> | .086<sup>***</sup> | .037 | .096<sup>***</sup> | -.058<sup>**</sup> | -.213<sup>***</sup> | -.129<sup>***</sup> | 1 |  |
| 14. Alcohol problems | -.060<sup>**</sup> | -.021 | .049<sup>*</sup> | -.043 | .061<sup>**</sup> | -.047<sup>*</sup> | .011 | -.058<sup>**</sup> | -.007 | .120<sup>***</sup> | .094<sup>***</sup> | .122<sup>***</sup> | -.032 | 1 |

\*\*\* p<0.001, \*\* p<0.01, \* p<0.05

The data were analysed using SPSS software (26.0 for Windows). P-values below 0.05 (two-tail) were considered statistically significant.

## Results

### Descriptive results

Because of high number of participants, distributions of FUNCT and COVFUNCT differed by background characteristics most often highly significantly (Table 1). The participants, who in the gender question selected the option “Other” (41, 2.1%) and “I do not want to tell” (30, 1.5%), reported lower FUNCT and COVFUNCT. Because these options might overlap, for further analyses, they were combined NBG (71, 3.6%).

The participants, who were older than 40 years, lived in marital relationship, were fulltime workers or worked at home and actively practiced physical exercise, reported the highest FUNCT and COVFUNCT scores, while use of mental care associated with low FUNCT and COVFUNCT.

FUNCT correlated with age, resilience, family and friend contacts, physical exercise, school success and close friends at age of 12 to 18. Loneliness, childhood adversities, depressive and anxiety symptoms and alcohol problems correlated negatively with FUNCT (Table 2). COVFUNCT correlated positively with age, resilience, family contacts, physical exercise, and negatively with loneliness, friend contacts, childhood adversities, and depressive and anxiety symptoms (Table 2).

The associations of gender, age, and work status with FUNCT and COVFUNCT are shown in Figure 1. After age of 41 FUNCT scores decreased with decreasing age, while COVFUNCT decreased after age of 51 linearly with decreasing age. Difference between BG and NBG was significant in both FUNCT (p<0.001) and COVFUNCT (p<0.001), while difference between university personnel and students was significant in FUNCT (p<0.001) but not in COVFUNCT (p=0.061).

**Figure 1.**
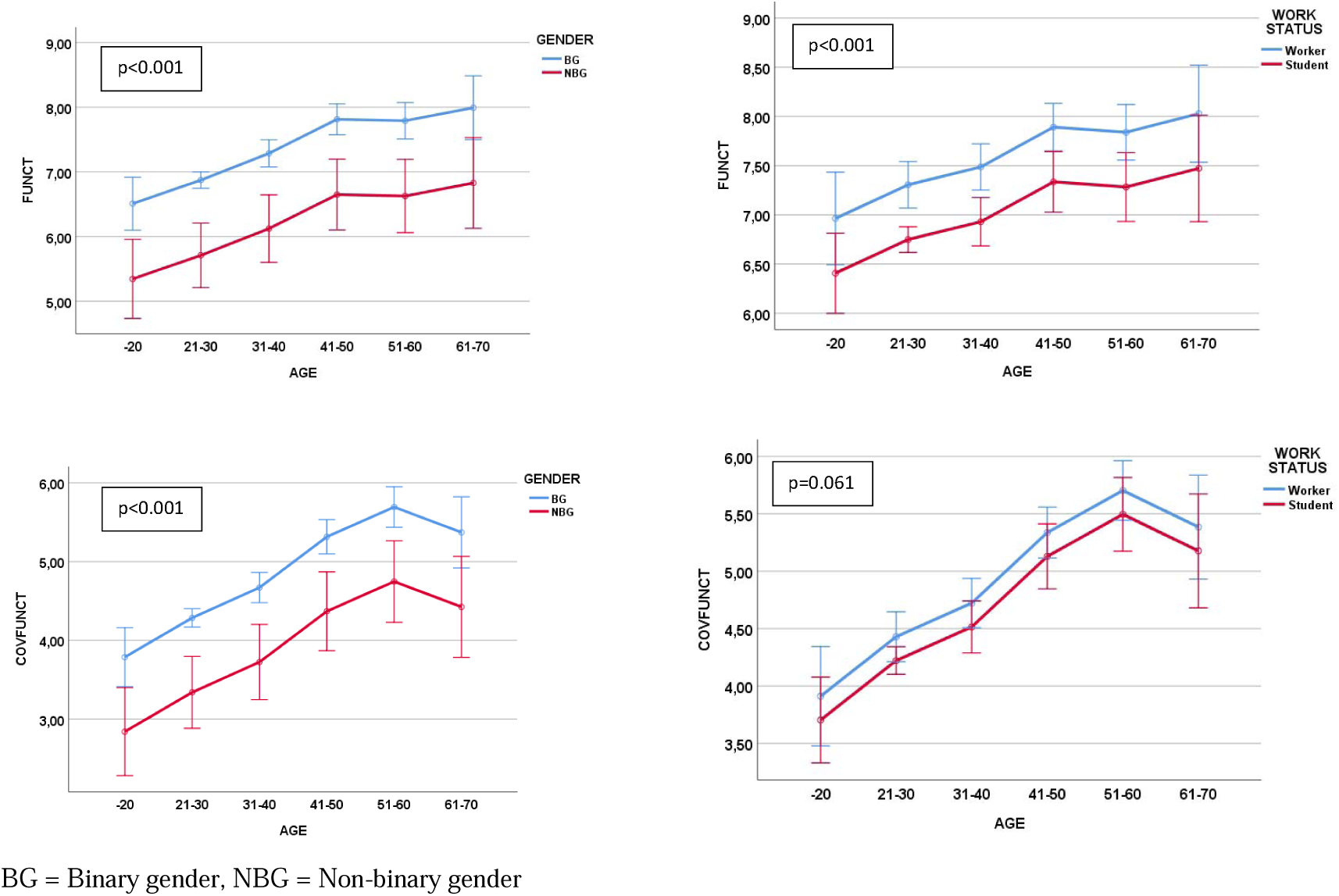
Functioning (FUNCT, 0-10) and effect of Covid-19 on functioning (COVFUNCT; 0-10) by gender, work status and age.

### Analyses of variance

In ANOVA, working or studying, active physical exercise, good resilience and good school performance in adolescence associated with good FUNCT, while being NBG, loneliness, received mental care during past 6 months or earlier, and childhood adversities associated significantly with poor FUNCT (Table 3 A). In other words, when the effects of other factors were taken into account, NBGs had lower FUNCT than BFs and BMs. However, there was no significant difference (p=0.060) between BFs and BMs. The employed participants had better FUNCT than students (p<0.001). Differences between daily and weekly physical exercise were also statistically significant (p=0.001). When depressive and anxiety symptoms were entered into the model, BF subjects had better FUNCT than BMs (p=0.049) and NBGs (0.014) (Table 3 B). There was no difference between BMs and NBGs. Because depressive symptoms associated strongly with loneliness, it became excluded from the model.

**Table 3.** ANOVA for functioning (A and B) and for the effect of covid-19 on functioning (C and D).

|  | A. Functioning |  |  |  | B. Functioning* |  |  |  | C. Effect of covid-19 on functioning |  |  |  | D. Effect of covid-19 on functioning* |  |  |  |
| --- | --- | --- | --- | --- | --- | --- | --- | --- | --- | --- | --- | --- | --- | --- | --- | --- |
|  | B | p | CI95% |  | B | p | CI95% |  | B | p | CI95% |  | B | p | CI95% |  |
| Intercept | 4.63 | <0.001 | 3.84 | 5.42 | 8.44 | <0.001 | 7.73 | 9.15 | 4.48 | <0.001 | 3.73 | 5.24 | 6.31 | <0.001 | 5.59 | 7.04 |
| Gender |  |  |  |  |  |  |  |  |  |  |  |  |  |  |  |  |
| Female | 0.74 | <0.001 | 0.32 | 1.15 | 0.44 | 0.014 | 0.09 | 0.79 | 0.93 | <0.001 | 0.51 | 1.35 | 0.68 | 0.001 | 0.28 | 1.08 |
| Male | 0.56 | 0.011 | 0.13 | 1.00 | 0.29 | 0.129 | -0.08 | 0.65 | 0.68 | 0.003 | 0.23 | 1.12 | 0.49 | 0.023 | 0.07 | 0.91 |
| Other | ref. |  |  |  | ref. |  |  |  | ref. |  |  |  | ref. |  |  |  |
| Work status |  |  |  |  |  |  |  |  |  |  |  |  |  |  |  |  |
| At work | 1.73 | <0.001 | 1.33 | 2.13 | 1.25 | <0.001 | 0.91 | 1.59 | 0.47 | 0.023 | 0.07 | 0.88 | 0.26 | 0.187 | -0.13 | 0.64 |
| Studying | 1.36 | <0.001 | 0.97 | 1.75 | 1.11 | <0.001 | 0.78 | 1.44 | 0.02 | 0.915 | -0.38 | 0.42 | -0.11 | 0.560 | -0.49 | 0.26 |
| Other | ref. |  |  |  | ref. |  |  |  | ref. |  |  |  | ref. |  |  |  |
| Physical exercise |  |  |  |  |  |  |  |  |  |  |  |  |  |  |  |  |
| Daily | 0.96 | <0.001 | 0.62 | 1.29 | 0.48 | 0.001 | 0.19 | 0.77 | 0.88 | <0.001 | 0.53 | 1.22 | 0.60 | <0.001 | 0.26 | 0.93 |
| Weekly | 0.45 | <0.001 | 0.26 | 0.64 | 0.23 | 0.005 | 0.07 | 0.39 | 0.11 | 0.257 | -0.08 | 0.31 | -0.03 | 0.767 | -0.21 | 0.16 |
| Less often | ref. |  |  |  | ref. |  |  |  | ref. |  |  |  | ref. |  |  |  |
| Meeting friends |  |  |  |  |  |  |  |  | -0.15 | <0.001 | -0.22 | -0.08 | -0.11 | 0.001 | -0.18 | -0.05 |
| Resilience | 0.11 | <0.001 | 0.09 | 0.13 | 0.03 | <0.001 | 0.02 |  | 0.05 | 0.09 | <0.001 | 0.06 | 0.11 | 0.04 | <0.001 | 0.02 |
| Loneliness | -0.72 | <0.001 | -0.85 | -0.58 |  |  |  |  | -0.56 | <0.001 | -0.70 | -0.41 |  |  |  |  |
| Mental care. 6 months |  |  |  |  |  |  |  |  |  |  |  |  |  |  |  |  |
| Yes | -0.77 | <0.001 | -0.99 | -0.55 | -0.43 | <0.001 | -0.62 | -0.24 | -0.41 | <0.001 | -0.61 | -0.22 |  |  |  |  |
| No | ref. |  |  |  | ref. |  |  |  | ref. |  |  |  |  |  |  |  |
| Mental care. ever |  |  |  |  |  |  |  |  |  |  |  |  |  |  |  |  |
| Yes | -0.41 | <0.001 | -0.60 | -0.22 | -0.30 | <0.001 | -0.46 | -0.14 |  |  |  |  |  |  |  |  |
| No | ref. |  |  |  | ref. |  |  |  |  |  |  |  |  |  |  |  |
| School success | 0.07 | 0.003 | 0.02 | 0.11 | 0.06 | 0.002 | 0.02 | 0.10 |  |  |  |  |  |  |  |  |
| Friends |  |  |  |  |  |  |  |  | -0.05 | 0.001 | -0.08 | -0.02 | -0.04 | 0.009 | -0.07 | -0.01 |
| Childhood adversities | -0.01 | 0.001 | -0.02 | -0.01 |  |  |  |  |  |  |  |  |  |  |  |  |
| Depressive symptoms |  |  |  |  | -0.17 | <0.001 | -0.18 | -0.15 |  |  |  |  | -0.11 | <0.001 | -0.12 | -0.10 |
| Anxiety symptoms |  |  |  |  | -0.01 | 0.001 | -0.02 | -0.01 |  |  |  |  |  |  |  |  |

\* Effects of depressive (DEPS) and anxiety (ANX) symptoms controlled.

In the next ANOVA, employment, daily active physical exercise and good resilience associated positively with COVFUNCT, while being NBG, meeting friends, loneliness, mental care during last six months and having close friends at age 12 to 18 associated negatively with COVFUNCT (Table 3 C). Negative associations of adolescence and current contacts with friends may indicate individuals’ social character and their effort to compensate the detrimental effect of Covid-19 on their functioning. The difference between BFs and BMs was non-significant (p=0.061). The participants being at work reported higher COVFUNCT scores than students (p<0.001).

When depressive and anxiety symptoms were entered into the model, NBGs and students (compared with being at work, p<0.001), meeting friends and having close friends at age 12 to 18 associated with negative, while daily active physical exercise and good resilience associated with positive COVFUNCT (Table 3D). BFs reported less negative COVFUNCT than BMs (p=0.024), and the participants being at work reported less negative COVFUNCT than students (p<0.001).

## Discussion

### Covid-19 effects of functioning and age

In line with previous studies (Prout et al, 2020; Guldager et al., 2021), age associated strongly with both FUNCT and COVFUNCT (see Figure 1). However, in multivariate analyses, because age correlated also with many other factors, associating more strongly with FUNCT and COVFUNCT, it was excluded from the ANOVA models. There was an interesting difference between FUNCT and COVFUNCT. The former decreased after age 41, while the later after age 51. This was caused by the fact, that the participants of age 51 to 60 reported the most positive changes in functioning during Covid-19 pandemic than other age groups (Figure 1).

During the Covid-19 pandemic, the age of 40 appeared to be a turning point among people in a university community. Older than that reported mainly positive (mean >5; range 0-10) COVFUNCT and they could keep their FUNCT on a reasonable level, while younger than that reported mainly negative (mean <5; rage 0-10) COVFUNCT and their FUNCT strongly decreased with decreasing age.

It is possible that in the university community, people older than 40 years represents a partly selected group in relation to the factors, such like resilience, stable work situation and relationships, supporting capability to resist the stress caused by the Covid-19 pandemic and restrictive measures related to it. Additionally, the age-related time (about 18 months) that the Covid-19 lockdown took from young people’s life was proportionally longer and therefore heavier than the same time in older people’s life. It is also probable, that the university lockdown measures (remote education and banned gatherings get-together) were targeted most heavily at young people, students in particular, who had been used to, and consequently more dependent on contact teaching and social contacts with friends than older people and personnel.

### Covid-19 effects of functioning and gender

In the present study, gender was considered in three categories: binary females, binary males, and reported non-binary gender. Combination of individual who selected the gender options: “Other” and “I do not want to tell” was argued by the fact that they both differed from females and males regarding psychic symptoms and FUNCT and COVFUNCT. The non-binary gender is an umbrella term including a variety of gender identities like genderqueer, genderless, gender-neutral, trigender, agender, third gender, two-spirit, and bigender (Chew et al., 2020). The prevalence (3.6%) of NBG received in the present study is small but comparable with the estimates reported in previous studies (Goodman et al., 2019; Chew et al., 2020; Zhang et al., 2020a).

Compared with BG participants, NBG participants had experienced more stressful events during their life cycle. Their parents were more often divorced, they reported a higher number of childhood adversities and had often used mental care services. Additionally, compared with BFs, their school performance had been lower and they had had fewer close friends in adolescence. At the time of the present survey, university lockdown for preventing Covid-19 viral infections had sustained for about a year and a half. At this stage, NBGs reported more loneliness, fewer contacts with family members and, in line with other studies (Prout et al., 2020; de Graaf et al., 2021; Schmits et al., 2021) more psychic stress symptoms, than BGs did, and their FUNCT was poorer than that of BGs although effects of other factors were controlled. Additionally, NBGs reported also more negative COVFUNCT than BFs and BMs did, indicating that they also suffered more from the stress caused by Covid-19 pandemic.

These findings, together with those regarding low resilience, support the view that NBG individuals were more vulnerable to the stress caused by the Covid-19 pandemic and its consequences than BG participants, suffered from increased psychic symptoms and consequently, decreased FUNCT. Non-binary individuals represent a disadvantageous, marginalised minority, being at risk of victimisation and discrimination stress (Rutherford et al. 2021; Schmits et al., 2021). Thus, it is understandable that additional social stress related to Covid-19 pandemic had so detrimental effect on their every-day FUNCT and that NBG individuals may need a special attention and support during ongoing and future pandemics. Differences in FUNCT between BGs were small because the factors associating with FUNCT had opposite effects: compared with BMs, BFs reported better school performance and more close friends at adolescence, and more contacts with family members and friends, but more childhood adversities, more loneliness and lower resilience. Although, BFs reported more received mental care and psychic symptoms, as also found by other researchers (Elmer et al., 2020; Guldager et al., 2021; Schmits et al., 2021), their FUNCT was at least as good as that of BMs suggesting that occurrence of psychic symptoms alone does not always mean decrease of FUNCT.

### Covid-19 and student status

Being at work, having good resilience and performing daily physical exercise associated consistently with good FUNCT and less negative COVFUNCT. Participants working at university community can be considered as a reference group for students whose FUNCT was clearly poorer than that of employed. In previous studies, students have reported worries related to Covid-19 pandemic, increase of workload, concern on academic performance, social isolation and loneliness and depression and anxiety (Elmer et al., 2020; Son et al., 2020; Wang et al., 2020; Guldager et al., 2021). In the present study, students reported lower resilience, more loneliness and psychic symptoms than employed persons did (not shown in analyses). However, when the effects of these factors were controlled, students still had lower FUNCT and they reported more negative effect of Covid-19 on their FUNCT than employed persons. Students’ life situation was not as established and their future prospects as certain as employed personnel though a great deal of personnel were temporary workers. Therefore, they had not capacity to resist the stress caused by Covid-19 and to keep their FUNCT at the pandemic preceding level during Covid-19 pandemic.

Interestingly, in ANOVA (Table 3C and 3D), meeting friends associated negatively with COVFUNT, but only in students, indicating that those of students who reported most negative COVFUNT met most often their friends. Possibly, they tried to compensate their decreased FUNCT by meeting friends at the time when meeting friends was strongly restricted.

### Covid-19, functioning and physical exercise

Covid-19 pandemic and related social aspects had strong negative effects on people’s physical activity. Therefore, encouraging people to move and take physical exercise is important for their functioning (Burtscher et al., 2020; Lesser & Nienhuis, 2020). In line with a previous study (Schmits et al., 2021), active physical exercise associated with good FUNCT and less negative COVFUNT. There was an interesting difference between FUNCT and COVFUNT: at least weekly exercise “at least half an hour at the time so that at least mildly get out of breath and sweat” associated with good FUNCT, while daily exercise of so much effort was required for positive COVFUNCT. A more detailed comparison (not shown) revealed, that with students, daily exercise produced mean COVFUNCT score 5.0 (range 0-10; SD 2.1), while the comparable COVFUNCT score for employed personnel was 6.3 (range 0-10; 2.2). The difference was significant (p=0.001) indicating that for preventing negative COVFUNCT, students had to do physical exercise much more than personnel, and with daily exercise they could just reach the COVFUNCT level “no effect”, while employed university personnel with same effort could improve their FUNCT.

### Covid-19 and resilience

Resilience refers to the construct involving the maintenance of, or return to, positive mental health following adversity by using a collection of multiple internal (personal characteristics or strengths) and external (qualities of wider family, social, and community environments) resilience protective factors (assets and resources) that enable an individual to thrive and to overcome disadvantage or adversity (Dray et al., 2017).

In line with previous studies (Ran et al., 2020; Hou et al., 2021a), low resilience associated with anxiety and depression symptoms. Furthermore, good resilience, measured by two VAS questions regarding recovery after an adversity and length of bothering after a failure, associated very strongly with actual good FUNCT and withstood Covid-19’s negative effects on individuals’ FUNCT. Among students, emotional resilience was positively correlated with learning management skills, and positive emotional ability predicted learning management skills (Zhang et al., 2020b).

In a population study, during the Covid-19 lockdown, average resilience was lower than published norms. However, it was greater among those, who exercised more and perceived more social support from family, friends, and significant others, indicating that psychological resilience in the face of the pandemic was related to modifiable factors (Killgore et al., 2020). In the present study, when many of these resilience-supporting factors were controlled, good resilience still associated positively with every day FUNCT and withstood Covid-19’s negative effects on FUNCT. This indicates to a personal strength to resist negative effects of the pandemic.

### Covid-19, functioning and childhood adversities

In the present study, childhood adversities associated strongly with reduced FUNCT in adulthood. In ANOVA, their significant association with negative effects of Covid-19 on FUNCT became explained by other factors. In line with other studies (Fryers & Burgha, 2013; Kessler at al., 2010; Salokangas et al., 2016), childhood adversities associated strongly with anxiety and depression symptoms, and when their effects on FUNCT were taken into account, childhood adversities became excluded from the ANOVA model. Most probably, the effect of childhood adversities on FUNCT was mediated via anxiety and depression symptoms.

### School success

Education, both compulsory schooling and post-compulsory schooling level, has an important causal effect in explaining differences in many adult outcomes including employment and health (Conti et al., 2010). It was interesting that although in the present study adolescence school success did not correlate very strongly with adult FUNCT, its positive effect on FUNCT remained significant in ANOVA indicating that school success is an important predictor for adult FUNCT even in a population with high academic education. Probably, adolescent school success indicates a good self-confidence – possibly at least partly developed within school success – that act as a longstanding buffer against the exceptional stress situation caused by the Covid-19 pandemic. It was not related to resilience or to reported effects of the pandemic.

### Loneliness

In line with meta-analyses (Loades et al., 2020; Gorenko et al., 2021), loneliness correlated strongly with depression and anxiety. In a review and meta-analysis, Covid-19 lockdown associated with anxiety and depression, but in multivariate analysis not with loneliness (Prati & Mancini, 2021). In accordance with the present study, taking into account the effects of anxiety and depression symptoms, loneliness was excluded from the ANOVA model, indicating that regarding FUNCT, psychic symptoms are more important than subjective feeling of loneliness that is actually one component of depression. Based on their findings Prati and Mancini (2021) suggest that lockdowns do not have uniformly detrimental effects on mental health and that most people are psychologically resilient to these effects.

### Advantages and limitations

In addition to important and actual findings related to the Covid-19 pandemic, some strengths as well as limitations should be discussed. The low response rate (7.4%) clearly limits representativeness of the results. On the other hand, the sample size became large enough for studying also small groups of participants and possible associations between various factors and functioning.

The survey was not very long but it included also sensitive questions, which may have reduced individuals’ willingness to response. In client satisfaction surveys with no incentives, response rate often remains on the level of 10% or under (PeoplePulse, 2021). During the Covid-19 pandemic, the university students and personnel received several other surveys, thus it is probable that they were tired to response to a new survey. Additionally, the fact that this survey was carried out in May, when the term was near to end, may have affected low response rate.

The study focused on people of university community, who do not represent the general population. On the other hand, the study sample represents a quite homogenous population, which faced equal and long-lasting Covid-19 lockdown with its consequences, when the differences in FUNCT between sub-groups of the sample, were mainly due to sub-group qualities than to different impacts of the pandemic.

The question on gender included only four options (Female, Male, Other, I do not wish to tell) and all except reported females and males were combined to one group of non-binary gender. This is heterogeneous group including several types of genderqueers and individuals who may be afraid that they can be traced from their responses. Therefore, results related to gender should be regarded provisionally, not conclusive; more detailed studies are needed.

## Data Availability

According to the decission of the ethical committee data are available only to the authors, not others.

